# Longitudinal gut microbial signals are associated with weight loss: insights from a digital therapeutics program

**DOI:** 10.1101/2023.01.04.22284035

**Authors:** Shreyas V. Kumbhare, Inti Pedroso, Bharat Joshi, Karthik M. Muthukumar, Santosh K. Saravanan, Carmel Irudayanathan, Gursimran S. Kochhar, Parambir S. Dulai, Ranjan Sinha, Daniel E. Almonacid

## Abstract

Obesity is a significant health problem due to its profound health deteriorating effects and high costs for healthcare systems. There exist lifestyle and pharmacological interventions available to prevent and reverse obesity; however, at the population level, these have shown to be insufficient, and we continue to see a worldwide increase in obesity prevalence. The gut microbiome has been shown to influence the susceptibility to weight gain and difficulty in losing weight and to be associated with successful long-term weight loss. Therefore, multiple studies have suggested that obesity interventions should consider the gut microbiome as a primary target through an improved diet and a crucial endpoint to monitor. However, there is a paucity of evidence regarding how to tailor the diet for an individual’s microbiome and what changes are expected to occur due to successful weight management. Digital therapeutics solutions have emerged as an exciting alternative to increase population access, reduce costs, and have the potential to accompany individuals on their health-promoting journey closely. Digbi Health has developed a dietary and lifestyle program to achieve weight loss that effectively reduces weight and improves diverse health outcomes by prioritizing and personalizing food ingredients to match an individual’s genetic profile and nurture the gut microbiome. In this study, we analyze the weight loss pattern and microbiome profile of 103 individuals to identify the effects of the weight loss program on the gut microbiome between their baseline and follow-up samples. We found that 80% of individuals lost weight during the study.

Analysis of their gut microbiome identified genera, functional pathways, and microbial communities associated with BMI changes and dietary and lifestyle program. The microbial genera and functional pathways associated with a reduction in BMI during the study include several previously reported in the literature, including *Akkermansia, Christensenella*, Oscillospiraceae, *Alistipes*, and *Sutterella*, short-chain fatty acid (SCFA) production and degradation of simple sugars like arabinose, sucrose, and melibiose. Network analysis identified two microbiome communities associated with BMI, one of which also significantly responded to the weight loss program, which includes multiple known associations with BMI and obesity. Our findings provide additional evidence for using the gut microbiome as an endpoint of weight loss program and highlight how it positively impacts the gut microbiome, with significant parallels in weight loss and health outcomes. These results provide additional evidence for known microbiome biomarkers of obesity and highlight new ones that warrant further research.

## Introduction

Obesity and related comorbidities are a public health priority, with rising costs for private and public health systems. Despite continuous efforts, most interventions fail at a population level to achieve effective long-term results. Digital therapeutic interventions have become increasingly popular for weight management and other health-related goals^1,2^. These interventions offer a convenient and cost-effective way for individuals to access personalized support and guidance for achieving their weight loss goals but differ in efficacy.

Obesity is a complex disorder influenced by genetic, environmental, and lifestyle factors. Studies in the past decade have unveiled the critical role of the gut microbiome not just in the development of obesity but also have highlighted its contribution to the reversal of metabolic disorders in general^3–6^. The human gut microbiome is a complex and dynamic community of microorganisms essential for maintaining human health^7^, and it is influenced by various factors, including diet and lifestyle^8–10^. The human gut microbiome plays a critical role in the metabolism of nutrients and other substances and can produce a wide range of beneficial and harmful metabolites^11^. Changes in the taxonomic composition of the gut microbiome can therefore affect the balance of microbial metabolic pathways, leading to changes in the types and amounts of metabolites produced, with important implications for human health.

In recent years, there has been increasing interest in studying the longitudinal changes in the gut microbiome over time as a response to different interventions and their relationship to various health outcomes, including body mass index (BMI)^12^. Research studies have shown that other diets, including low-fat and high-fat, can alter the gut microbiome in obese individuals^13,14^. For example, a high-fat diet may increase the abundance of certain bacteria associated with obesity. In contrast, a low-fat diet may increase the abundance of bacteria associated with leanness^5,15,16^. A few reports have shown differences in the gut microbiome taxonomic composition, diversity, and metabolic pathways encoded in individuals with different BMIs^12,17–19^, and that different dietary patterns result in distinct patterns of microbial taxa^8^. Although it is evident from studies that dietary interventions, such as changing the types and amounts of food that are consumed, can affect the composition and function of the gut microbiome, it is not well understood how these differences evolve over time and whether they play a role in the development or reversal of obesity^12^.

Recent advances in genomics and microbiome research have provided insights into their role in weight regulation, allowing for the development of targeted dietary interventions for weight loss based on these factors^18,20–22^. Digbi Health has developed a digital therapeutics program based on the participants’ multi-omic signals (genetics and gut microbiome), along with self-reported data and data from wearable devices. The outcomes of the weight loss program in different cohorts of individuals have been recently published, demonstrating its applicability not only in weight loss^23^, but also in reducing fasting blood glucose and HbA1c levels^24^, improving mental health symptoms^25,^ and functional gastrointestinal disorders^26^.

In this study, we aimed to investigate the effects of a personalized dietary and lifestyle weight loss program on the gut microbiome composition over time and its relationship to BMI. To understand the impact of this program on the gut microbiome, we performed longitudinal microbiome sampling at the beginning and approx at six months follow up. Our analysis focused on changes over time in the relative abundances of microbial taxa and their encoded metabolic pathways, alpha and beta diversity, and the structure of microbial co-abundance networks in response to the weight loss program.

## Results

### Cohort characteristics

The cohort enrolled in the weight loss program comprised 75.7% of females with an average age of 53.55 (Median 55.0; IQR: 44.5, 63.0). Almost 60% of individuals were suffering from FGIDs (at least one self-reported functional gastrointestinal disorder: IBS, gassiness, bloating, constipation, diarrhea, or dyspepsia), while ∼86% had other comorbidities along with overweight or obese conditions at baseline (T0). 35% of individuals were on prescribed antidepressants or anxiolytics, while 14.6% were using recreational drugs at baseline (T0) during enrollment.

We noted a statistically significant weight loss from baseline until T2, with around 80% of individuals losing weight. On average, the reduction in BMI from T0 to T2 was 2.57 BMI units (Median: 2.6; IQR 2.1, 2.6, p-value<0.001). There was a significant reduction (p-value<0.001, Wilcoxon Signed-rank test) in weight from T1 to T2 with an average decrease of 1.6 BMI units (Median 1.4 (IQR: 0.3, 2.7); see Table 1 and Figure 1A).

**Table 1.**
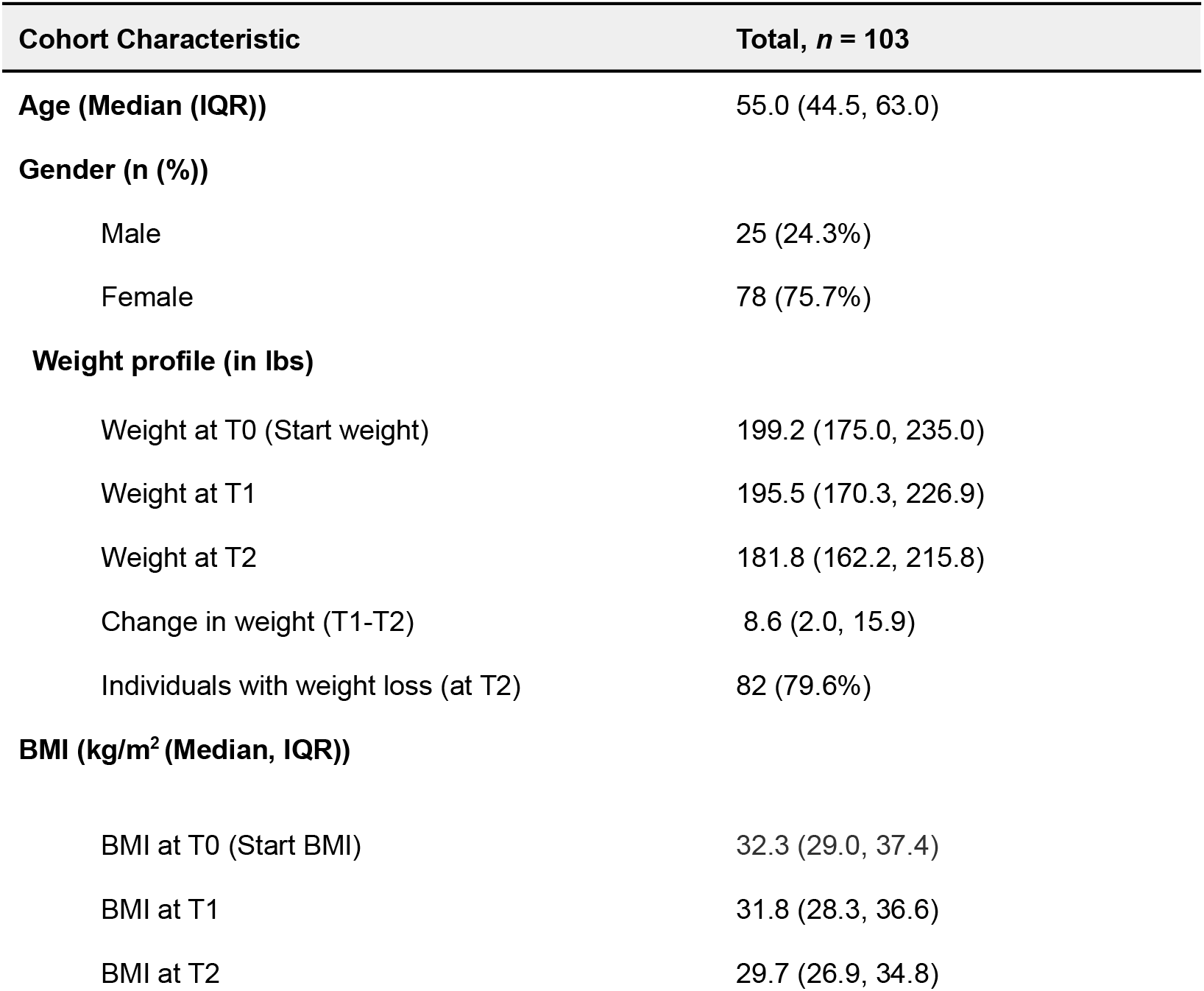
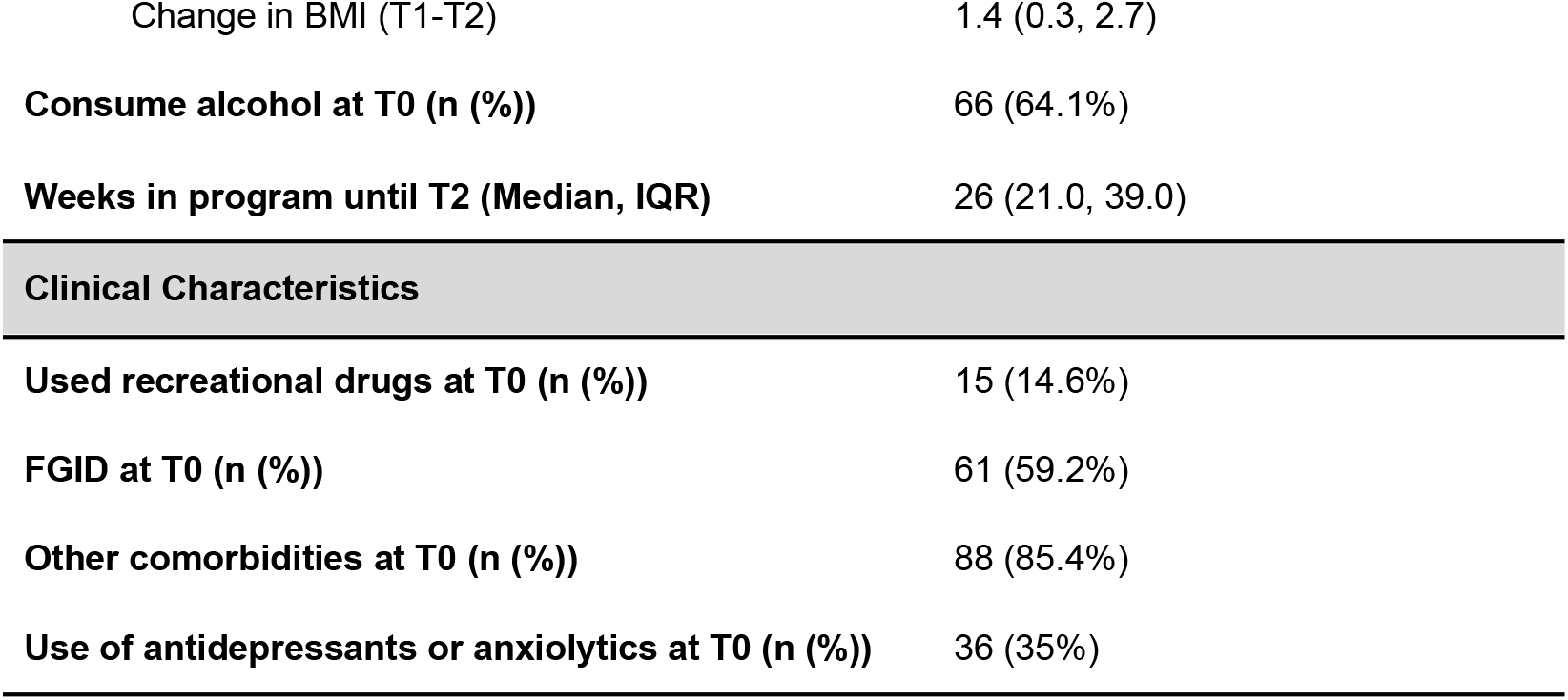
Study cohort and demographic characteristics. Values are expressed as median (IQR) and percentages of available data. FGID: Functional Gastro-Intestinal Disorders; BMI: Body Mass Index; T0: at Baseline (during enrollment), T1: Early phase, T2: Follow-up phase.

| Cohort Characteristic | Total, *n* = 103 |
| --- | --- |
| **Age (Median (IQR))** | 55.0 (44.5, 63.0) |
| **Gender (n (%))** |  |
| Male | 25 (24.3%) |
| Female | 78 (75.7%) |
| **Weight profile (in lbs)** |  |
| Weight at T0 (Start weight) | 199.2 (175.0, 235.0) |
| Weight at T1 | 195.5 (170.3, 226.9) |
| Weight at T2 | 181.8 (162.2, 215.8) |
| Change in weight (T1-T2) | 8.6 (2.0, 15.9) |
| Individuals with weight loss (at T2) | 82 (79.6%) |
| **BMI (kg/m<sup>2</sup> (Median, IQR))** |  |
| BMI at T0 (Start BMI) | 32.3 (29.0, 37.4) |
| BMI at T1 | 31.8 (28.3, 36.6) |
| BMI at T2 | 29.7 (26.9, 34.8) |
| Change in BMI (T1-T2) | 1.4 (0.3, 2.7) |
| Consume alcohol at T0 (n (%)) | 66 (64.1%) |
| Weeks in program until T2 (Median, IQR) | 26 (21.0, 39.0) |
| **Clinical Characteristics** |  |
| Used recreational drugs at T0 (n (%)) | 15 (14.6%) |
| FGID at T0 (n (%)) | 61 (59.2%) |
| Other comorbidities at T0 (n (%)) | 88 (85.4%) |
| Use of antidepressants or anxiolytics at T0 (n (%)) | 36 (35%) |

**Figure 1.**
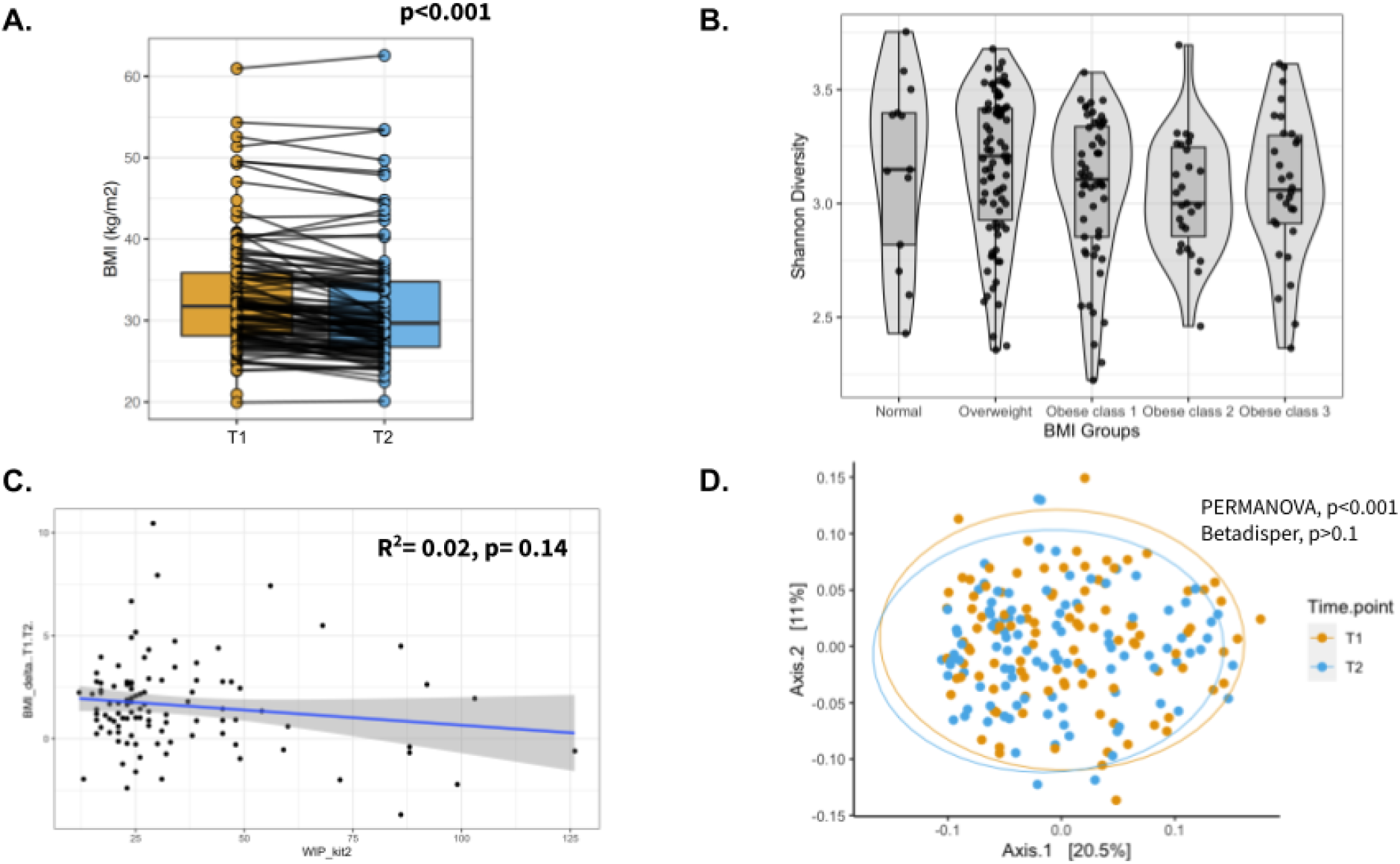
Weight loss after participating in the program and gut microbial composition. (A) Box-plot of BMI for the individuals at T1 and T2. Measurements of the same individual are linked with a line across the two-time points. p-value from Wilcoxon-signed rank test. (B) Box-plot displaying the Shannon diversity across different BMI groups. (C) Scatter plot showing the regression analysis of change in BMI (T2-T1) and Weeks in Program (WIP) until T2. (D) Principal Coordinate Analysis (PCoA) plot showing the beta diversity of the gut microbes across all individuals based on the Bray-Curtis dissimilarity. Ellipses represent 95% confidence regions. PERMANOVA analysis showed significant differences (shift) in overall diversity between two-time points (p-value < 0.001). PERMANOVA: Permutational Multivariate Analysis of Variance; T1: Early phase; T2: Follow-up phase.

Around 14.6% of individuals lost 3-5%, 34% lost 5-10%, and 28.2% lost more than 10% of body weight from baseline T0 to T2. Furthermore, 17.5% of individuals lost 3-5%, 31.1% lost 5-10%, and 14.6% lost more than 10% of body weight from T1 to T2. Although there was variation in the weeks in the program across the cohort until T2, it had no significant association with the amount of weight loss (p-value = 0.14) (Figure 1C). Additionally, we did not observe any significant influence of age, gender, BMI at T0, and use of anxiolytics and antidepressants at T0, except alcohol consumption at T0 on percentage body weight loss (Table S1).

#### BMI is associated with alpha and beta gut microbial diversity

We further implemented a multivariate association that showed that along with sampling time point (T1 vs. T2 PERMANOVA, p-value <0.001), Age (PERMANOVA, p-value <0.01), and Gender (PERMANOVA, p-value <0.01) were among the top drivers of overall gut microbial diversity (see Figures 1D, 2 and Table 2). Interestingly, the inter-individual variation, when compared between the genders (Male vs. Female), was significantly different (Betadipser test p-value = 0.01) and reduced at T2 (Figure 2A). Additionally, the consumption of prescribed antidepressants and anxiolytics and the consumption of alcohol at baseline (T0) were seen to contribute significantly to the microbiome variation. BMI had a significant association with the overall diversity (PERMANOVA, p-value = 0.04) and was not confounded by other variables, including age, gender, and time point, as evidenced by the interactions (Table 2 & Figure 2).

**Table 2.**
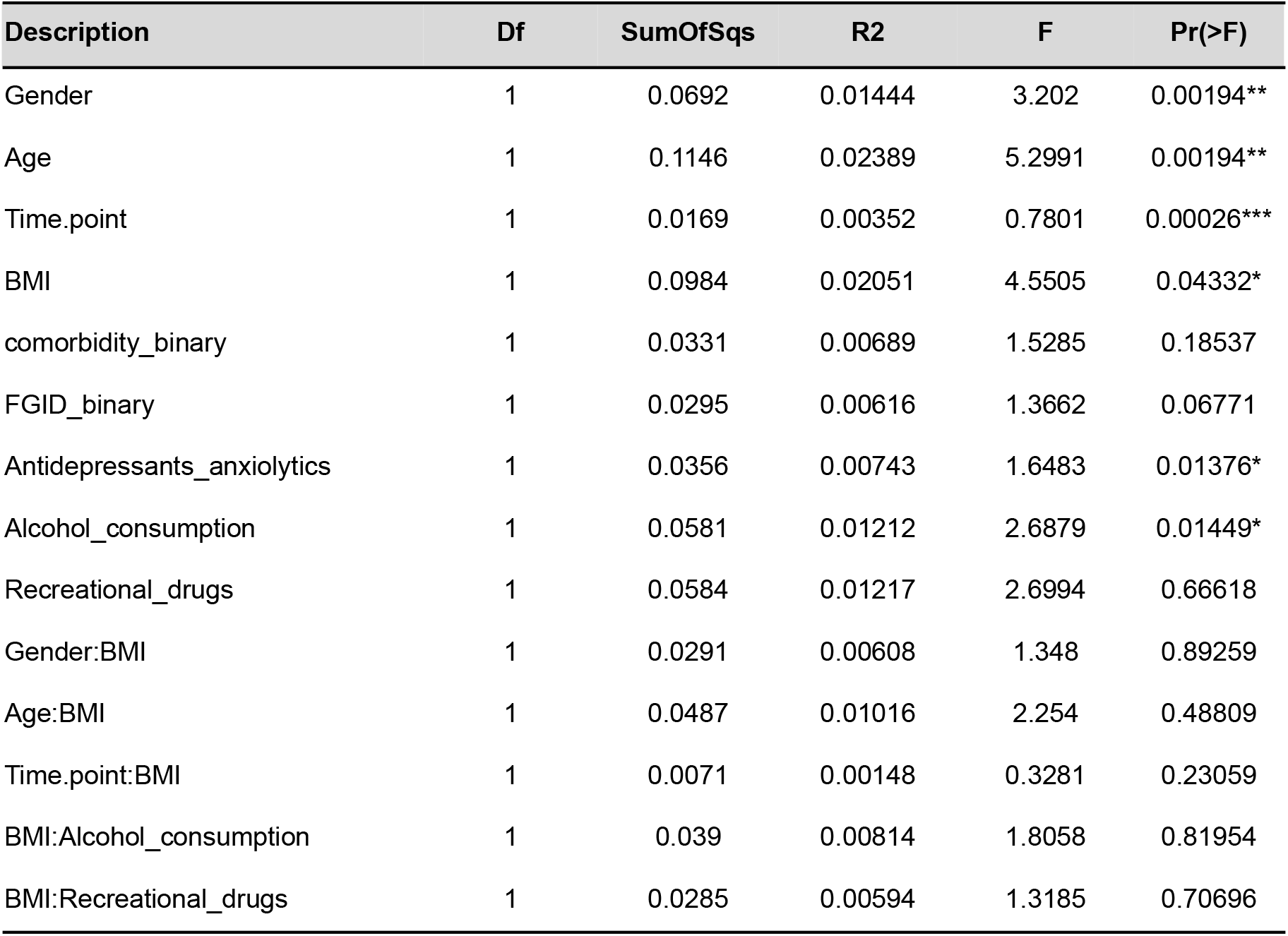
Results from multivariate analysis (PERMANOVA) showing an association between variables and gut microbial diversity. PERMANOVA: Permutational Multivariate Analysis of Variance.

**Figure 2.**
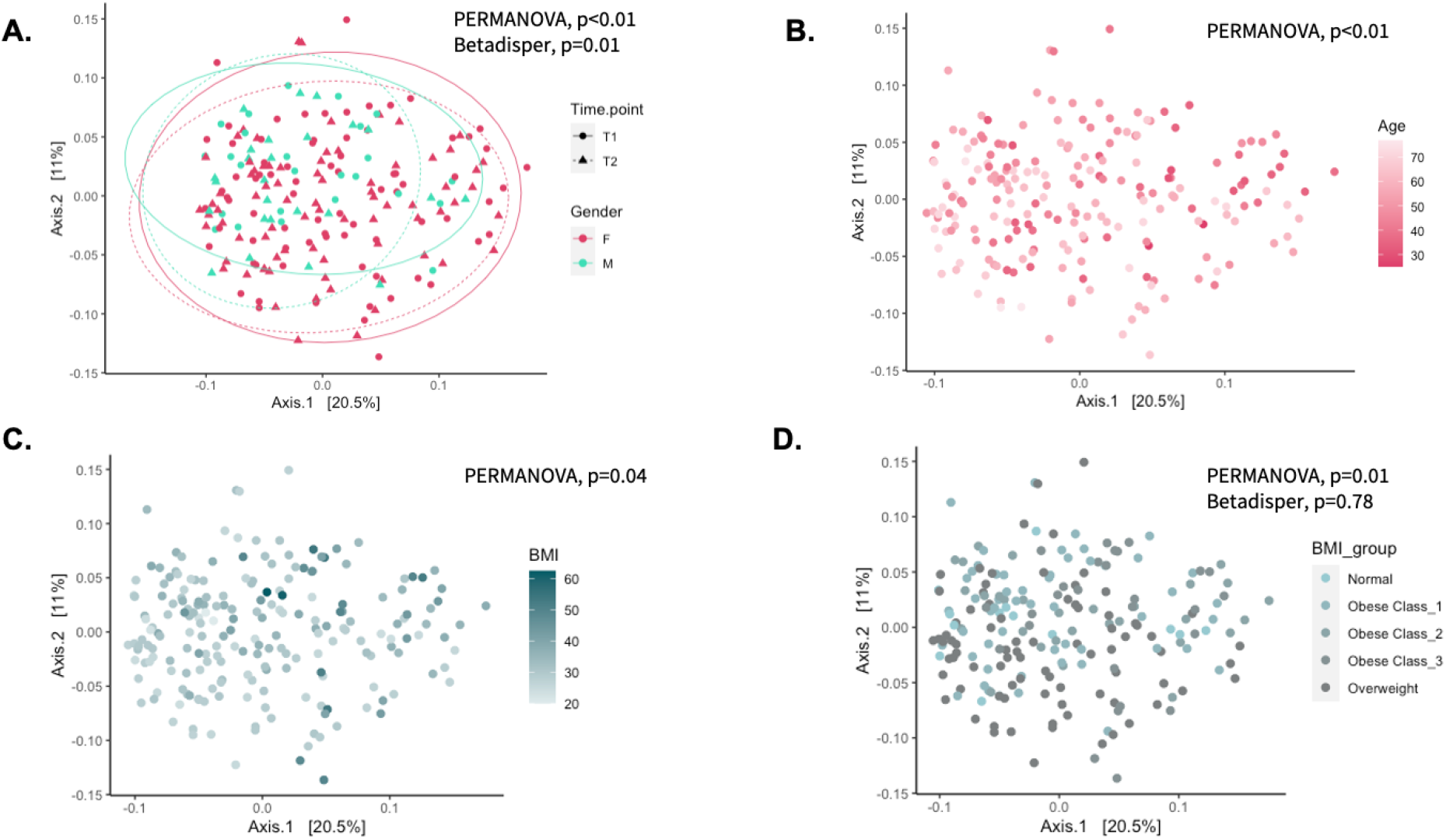
Association of Gender, Age and BMI with gut microbial diversity. Principal Coordinate Analysis (PCoA) plot showing the beta diversity of the gut microbes across all individuals based on the Bray-Curtis dissimilarity. Ellipses in panel-A represent 95% confidence regions, stratified by gender at each time point. PERMANOVA analysis showed significant differences (shift) in overall diversity (A) between gender (PERMANOVA: p-value < 0.01, Betadisper: Homogeneity of Variances test: p-value = 0.01), (B) with age (PERMANOVA: p-value < 0.01), (C) with BMI (PERMANOVA: p-value = 0.04) and (D) between BMI groups (PERMANOVA, p-value = 0.01, Betadisper: Homogeneity of Variances test: p-value = 0.78). PERMANOVA: Permutational Multivariate Analysis of Variance; T1: Early phase; T2: Follow-up phase.

We also tested the influence of these predictors on Shannon and Simpson’s diversity measures using linear mixed model analyses implemented on the GAMLSS regression method. We identified a significant positive association between Age and Shannon (p-value = 1.6×10^−4^) and Simpson’s (p-value = 2.9×10^−2^) diversity and a negative association between BMI and Shannon (p-value = 2.7×10^−3^) but not with Simpson’s (p-value = 0.39) diversity (Table S2).

### Gut microbial signals are associated with the successful weight loss

As evident from the PERMANOVA model, the time point in the program (T1: Early phase vs. T2: Follow-up phase) contributed to the variation in microbial diversity, indicating a shift in gut microbial diversity structure with weight loss program, and was independent of the variation explained by BMI. We applied a Generalized Additive Model for Location, Scale, and Shape (GAMLSS) with a zero-inflated beta (BEZI) family (GAMLSS-BEZI) regression model to analyze the association of variables with individual microbial taxa (at genus level) and predicted pathway’s abundance (see Tables S3 and S4). In total, 62 genera showed a significant association with BMI after correcting for multiple testing (FDR < 0.05). Of note, *Megasphaera, Cloacibacillus*, and members of the Lachnocpiraceae family (including *Lachnoclostridium*, unannotated genera such as Lachnospiraceae UCG-001 and Lachnospiraceae CAG-56) were associated with increasing BMI. While *Hydrogenoanaerobacterium, Solobacterium, Desulfovibrio, Phascolarctobacterium*, Christensenellaceae R7 group, and unannotated genera from Oscillospiraceae family (including UCG_002, UCG_005, NK4A214_group) were associated with lower BMI (see Table S3). In total, 36 genera were seen to change significantly at time point T2 (FDR < 0.05), indicating changes associated with the weight loss program. Of note, *Akkermansia, Anaerotruncus, Coprobacter*, and *Rothia* increased substantially at time point T2 (see Table S3).

We further investigated the 18 taxa associated with BMI and time points. Interestingly, 9 out of these 18 taxa had a pattern of association indicating that the weight loss program improved their abundance, i.e., negatively associated with BMI and increased at T2 or positively associated with BMI and decreased at T2. In particular, those associated with lower BMI, Clostridia vadinBB60 group, Christensenellaceae R7 group, *Enterobacter, Desulfovibrio*, and Oscillospiraceae UCG-002 were seen to be enriched at time point T2; on the other hand, genera associated with higher BMI *Agathobacter, Fenollaria, Lachnoclostrdium*, and *Roseburia* depleted substantially at T2 (See Figure 3A, Table S3). We also did notice a few taxa that increased at T2 and were associated with higher BMI, such as *Cloacibacillus, Enterohabdus, Megasphaera*, and *Sutterella*, while those that were associated with lower BMI and decreased at T2, such as *Eggerthella*, Erysipelatoclostridiaceae UCG-004, *Eubacterium ruminantium* group, *Phascolarctobacterium*, and *Solobacterium*.

**Figure 3.**
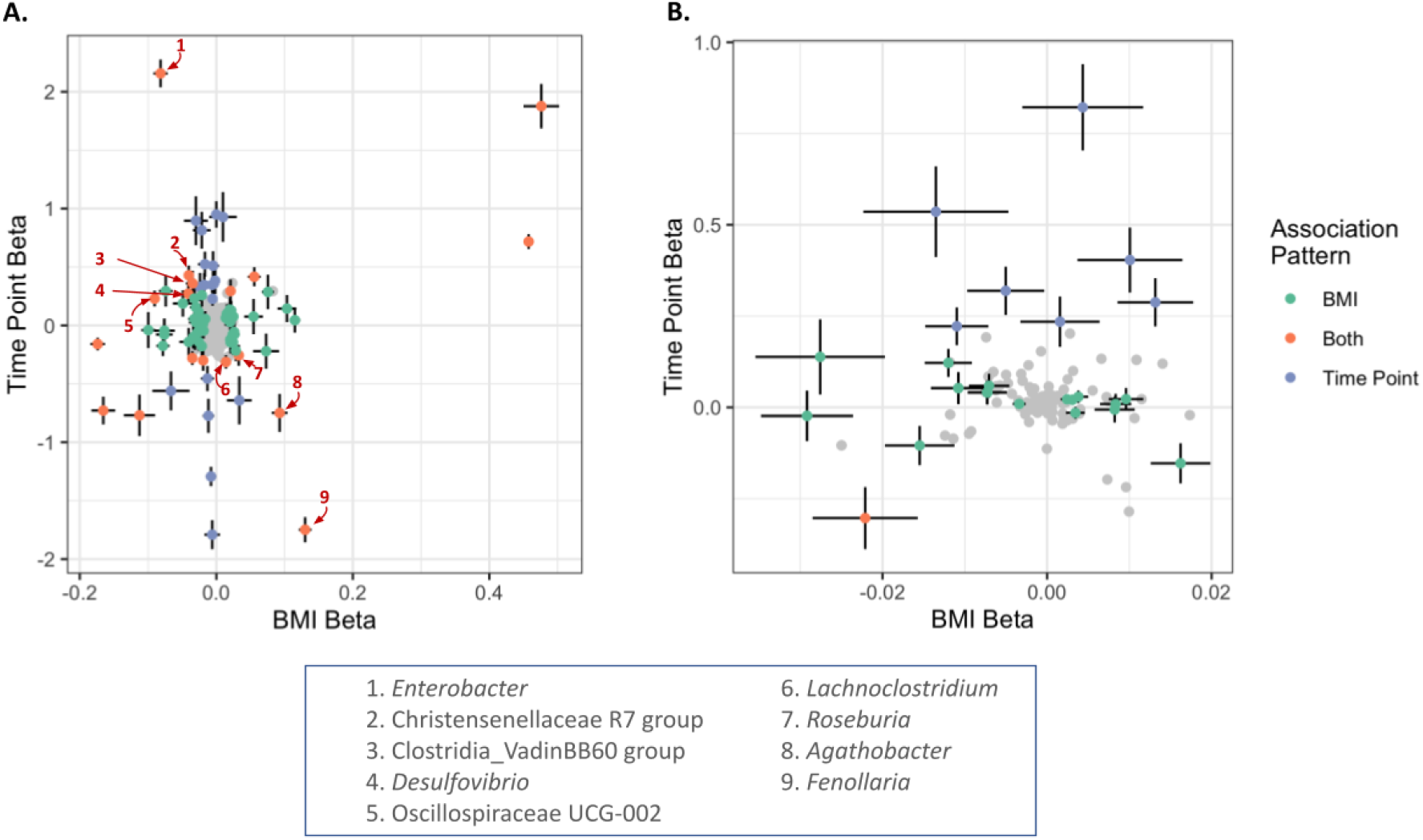
Microbial biomarkers associated with BMI and time point T2. Scatter plots of the regression effects size (coefficient) and its standard error of the association of each taxa (panel A) or functional pathway (panel B) with BMI (x-axis) and time point (y-axis) illustrating the association pattern of key taxa (A) and functional pathways (B). Points have been color-coded as indicated on the legend and associations were deemed significantly associated with an FDR < 0.05. Features highlighted with numbers and arrows in red in panel A were significantly associated with both BMI and time point T2 and were either negatively associated with BMI and enriched at T2 or positively associated with BMI and depleted at T2 as discussed in the main text.

Additionally, we tested the association with microbial functions. We found that the abundance of 21 pathway modules was significantly associated with BMI (FDR < 0.05), out of which pathways related to the degradation of simple sugars (carbohydrates) such as arabinose (MF0014), sucrose (MF0010) and melibiose (MF0009) were significantly associated with higher BMI (Table 3). On the other hand, biosynthesis of propionate (MGB054, MGB055, and MF0094), a Short Chain Fatty Acid (SCFA), GABA (γ-Aminobutyric acid) synthesis (MGB021), putrescine degradation (MF0082), degradation of amino acids such as lysine (MF0057), serine (MF0048) and phenylalanine (MF0024), and triacylglycerol degradation (MF0064) were negatively associated with higher BMI. The association between time point and predicted functions that reached significance after multiple testing corrections (FDR < 0.05) consisted of pathways related to nitric oxide metabolism (MGB026, MGB027, MGB028), inositol (MGB038) & GABA (MGB019) degradation and pyruvate dehydrogenase complex (MF0072), which were seen at higher abundance at T2. In contrast, the pathway associated with histamine synthesis (MGB009) was depleted at T2 (See Table 4). In line with the results for bacterial taxa, we observed a functional pathway, histamine synthesis, which was associated with BMI and time point (see Figure 3B).

**Table 3.** Association of gut microbial pathways and BMI.

| Module | Pathway name | Estimate | Std. Error | t value | p-value | fdr |
| --- | --- | --- | --- | --- | --- | --- |
| MF0082 | putrescine degradation | -0.0292 | 0.0056 | -5.2155 | 7.72E-07 | 1.55E-05 |
| MGB021 | GABA synthesis II | -0.0292 | 0.0056 | -5.2155 | 7.72E-07 | 1.55E-05 |
| MF0102 | sulfate reduction<br>(dissimilatory) | 0.0096 | 0.0021 | 4.5682 | 1.08E-05 | 0.0002 |
| MF0001 | arabinoxylan degradation | 0.0083 | 0.0019 | 4.3886 | 2.22E-05 | 0.0003 |
| MF0043 | cysteine<br>biosynthesis/homocysteine<br>degradation | 0.0162 | 0.0036 | 4.4733 | 1.67E-05 | 0.0007 |
| MF0048 | serine degradation | -0.0034 | 0.0008 | -4.2582 | 3.90E-05 | 0.0008 |
| MF0057 | lysine degradation I | -0.0120 | 0.0029 | -4.1826 | 5.00E-05 | 0.0014 |
| MF0064 | triacylglycerol degradation | -0.0155 | 0.0042 | -3.6639 | 0.0004 | 0.0115 |
| MF0098 | hydrogen metabolism | 0.0082 | 0.0024 | 3.3645 | 0.0010 | 0.0133 |
| MF0024 | phenylalanine degradation | -0.0276 | 0.0079 | -3.5038 | 0.0006 | 0.0173 |
| MGB009 | Histamine synthesis | -0.0221 | 0.0064 | -3.4594 | 0.0007 | 0.0216 |
| MF0095 | propionate production III | -0.0108 | 0.0033 | -3.2377 | 0.0015 | 0.0258 |
| MGB055 | Propionate synthesis III | -0.0108 | 0.0033 | -3.2377 | 0.0015 | 0.0258 |
| MF0094 | propionate production II | -0.0073 | 0.0024 | -3.0672 | 0.0026 | 0.0299 |
| MF0080 | lactate consumption II | -0.0073 | 0.0024 | -3.0672 | 0.0026 | 0.0299 |
| MGB054 | Propionate synthesis II | -0.0073 | 0.0024 | -3.0672 | 0.0026 | 0.0299 |
| MF0014 | arabinose degradation | 0.0038 | 0.0012 | 3.1532 | 0.0020 | 0.0327 |
| MF0002 | fructan degradation | 0.0031 | 0.0010 | 3.1750 | 0.0019 | 0.0333 |
| MF0010 | sucrose degradation I | 0.0024 | 0.0008 | 3.0777 | 0.0025 | 0.0455 |
| MF0103 | mucin degradation | -0.0071 | 0.0025 | -2.8751 | 0.0047 | 0.0487 |
| MF0009 | melibiose degradation | 0.0034 | 0.0011 | 3.0052 | 0.0031 | 0.0499 |

**Table 4.** Impact of the weight loss program on the gut microbial pathways at Time Point-2 (T2).

| Module | Pathway name | Estimate | Std. Error | t value | p-value | fdr |
| --- | --- | --- | --- | --- | --- | --- |
| MGB026 | Nitric oxide synthesis II<br>(nitrite reductase) | 0.8219 | 0.1182 | 6.9547 | 1.24E-10 | 2.20E-09 |
| MF0076 | 4-aminobutyrate<br>degradation | 0.3192 | 0.0661 | 4.8255 | 3.69E-06 | 0.0002 |
| MGB019 | GABA degradation | 0.4035 | 0.0890 | 4.5331 | 1.27E-05 | 0.0010 |
| MGB027 | Nitric oxide degradation<br>I (NO dioxygenase) | 0.2877 | 0.0661 | 4.3501 | 2.79E-05 | 0.0015 |
| MGB038 | Inositol degradation | 0.2220 | 0.0518 | 4.2866 | 3.35E-05 | 0.0018 |
| MGB028 | Nitric oxide degradation<br>II (NO reductase) | 0.5359 | 0.1243 | 4.3130 | 3.21E-05 | 0.0027 |
| MGB009 | Histamine synthesis | -0.3033 | 0.0850 | -3.5686 | 0.0005 | 0.0261 |
| MF0072 | Pyruvate<br>dehydrogenase complex | 0.2346 | 0.0690 | 3.4017 | 0.0009 | 0.0368 |

### Association of gut microbiome networks with BMI

Weighted network analyses identified five network modules as the study samples’ overall organizational structure of the gut microbiome (See Figure 4A, Tables 5 and 6). These network modules ranged in size between 13 to 20 taxa. We used the module’s eigenvector as a summary of the abundance of the taxa of each module and tested its statistical association with BMI and the change between T1 to T2. Module 2 was associated with BMI with a positive correlation (Sidak p-value = 0.037) (Figure 4B). Module 3 was associated with BMI with a negative correlation (Sidak p-value = 0.015) and also showed a change in the abundance pattern between T1 and T2 with a negative correlation (Sidak p-value = 0.024) (Figure 4C and 4D). We identified the taxa driving each module’s co-abundance pattern captured by the eigenvector. Figure S1 presents the correlation pattern between each module’s top 3 driving genera and the eigenvector. Several genera identified on univariate analyses are part of these network modules and are among those identified as key taxa within the modules. Table S5 lists the taxa associated with network modules 2 and 3 and their correlation with their module’s eigenvector.

**Table 5.** Association of gut microbial network modules and BMI.

| Module_name | Module Color | Estimate | Std. Error | df | t value | p-value | Sidak p-value |
| --- | --- | --- | --- | --- | --- | --- | --- |
| ME1 | turquoise | -0.001 | 0.0008 | 142.76 | -0.91 | 0.362 | 0.894 |
| ME2 | blue | 0.002 | 0.0008 | 126.06 | 2.72 | 0.007 | 0.037 |
| ME3 | brown | -0.001 | 0.0005 | 118.41 | -3.02 | 0.003 | 0.015 |
| ME4 | yellow | 0 | 0.0009 | 129.64 | 0.49 | 0.625 | 0.993 |
| ME5 | green | -0.002 | 0.0008 | 117.77 | -2.23 | 0.028 | 0.132 |

**Table 6.** Association of gut microbial network modules and Time point (T1 vs. T2).

| Module_name | Module Color | Estimate | Std. Error | df | t value | p-value | Sidak p-value |
| --- | --- | --- | --- | --- | --- | --- | --- |
| ME1 | turquoise | 0.0099 | 0.00476 | 100.94 | 2.078 | 0.04024 | 0.186 |
| ME2 | blue | -0.01261 | 0.00583 | 101.62 | -2.162 | 0.03294 | 0.154 |
| ME3 | brown | 0.01077 | 0.00374 | 99.18 | 2.88 | 0.00487 | 0.024 |
| ME4 | yellow | -0.00297 | 0.00595 | 101.52 | -0.499 | 0.61876 | 0.992 |
| ME5 | green | 0.004 | 0.00669 | 100.32 | 0.597 | 0.55157 | 0.982 |

**Figure 4.**
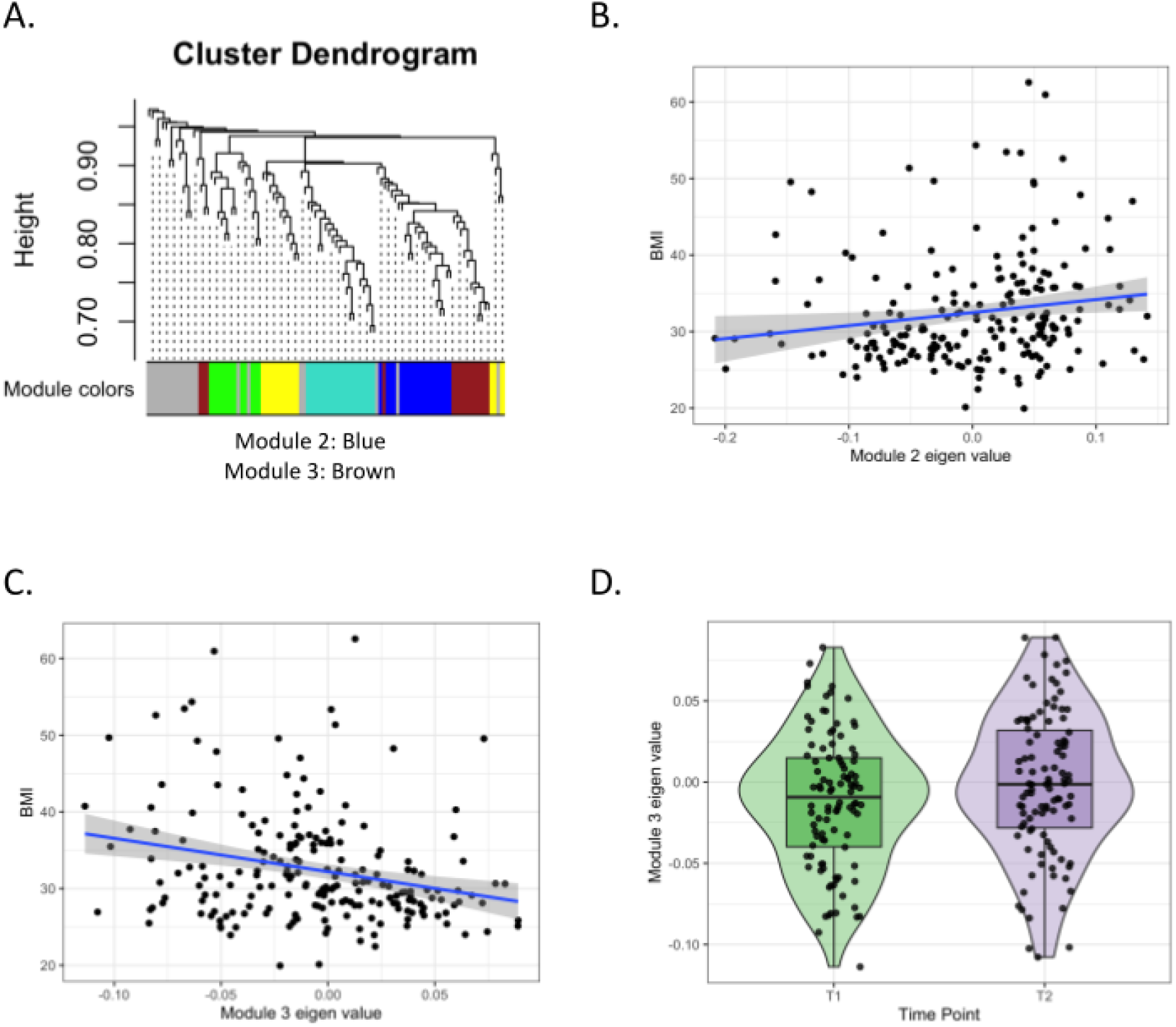
Association of gut microbial network modules and BMI. (A) The dendrogram presents the correlation among gut microbiome taxa abundance values on which each tip of the tree represents a genera and two genera are close to each other if they have a high signed bicor correlation value. Color blocks at the lower part of the figure represent the groups of taxa network modules identified as independent network modules. The relationship between colors and numbers is the following: turquoise = 1, blue = 2, brown = 3, yellow = 4, green = 5 and gray = 0. (B) and (C) Correlation between module 2 and 3 eigenvectors and BMI. The regression line (blue line and the grey area covers 1 standard deviation around the mean) represents the linear relationship between the two variables (D) box-plot displaying the module 3 eigenvector values across T1 and T2.

## Discussion

Our study analyzed a retrospective cohort of 103 individuals from their enrollment (T0) to study the effects of Digbi’s digital therapeutic obesity management and weight loss program on their BMI and gut microbiome (measured at T1 and T2) as primary endpoints. Overall we observed a significant weight loss across the study participants, with a mean weight decrease of 2.6 and 1.4 BMI units lost from T0 to T2 and T1 to T2, respectively (Table 1), corresponding to a mean of 7.4% and 4.9% body weight loss. Interestingly, we also observed a significant and beneficial impact on gut microbiome diversity, primarily associated with weight loss (Table 2 and S2). In individual association analyses, we identified gut microbiome biomarkers associated with weight loss and the sampling time point (T1 vs. T2) for bacterial genera (Table S3), bacterial functional pathways (Table S4), and bacterial community networks (Tables 5, 6, and S5). We did not find that the individual’s demographics or lifestyle factors analyzed influenced the weight loss (or gain) achieved between T1 and T2 (Table S1). This is in line with our previous scientific reports that show equitable results of Digbi’s weight loss program concerning the individual gender and age, among other socio-economic factors^25^.

Previous studies have reported a significant negative association between obesity or BMI and gut microbial diversity and a general reduction in gut microbiome diversity with chronic diseases^27–30^. Our results align with previous research and link decreasing BMI with an increase in alpha and its association with beta diversity (Tables 2 and S2). Interestingly, the change in beta diversity is ∼6x more strongly associated with BMI or age than with medication intake, alcohol consumption, or the time point studied (T1 vs. T2) (Table 2). This suggests that, among the variables tested, BMI is the main intra-individual factor driving the change in beta diversity between T1 and T2. Similarly, the alpha diversity analysis points toward BMI as the main factor influencing gut microbiome diversity during the studied period (Table S2). However, we must interpret these results with caution since individuals may have changed one or more additional determinants of health during the program. These may not have been included in our analyses nor reported through the digital therapeutics app. Therefore, we cannot rule out the systematic effect of other lifestyle changes or health conditions as partially responsible for improvements in microbiome diversity.

Through regression analyses, we identified the association between the abundance of 62 genera and the predicted abundance of 21 functional pathways with BMI, and 21 genera and nine functional pathways changed their abundance between T1 and T2 (Tables S3 and S4). We consider both results complementary since not all changes in the individual’s life during the program (between T1 and T2) that may affect the gut microbiome may be linked to BMI. Thus, we used the intersection between these two sets of results to identify genera and functional pathways that are physiological correlates of the BMI changes between T1 and T2. Eighteen genera were associated with BMI and T1 vs. T2 differences in abundance (Table S3), and several had been previously associated with BMI or obesity. *Christensenella* and Oscillospiraceae have been previously associated with BMI in humans. In particular, Oscillospiraceae are highly heritable in twin studies, and its abundance is possibly associated with that of *Christensenella*^31^. We observed a similar direction of association between these genera and BMI, and their abundances were positively correlated in our data and were part of a gut microbiome community network associated with BMI (Tables S3 and S5).

Amongst the predicted bacterial pathways, we observed a significant association of simple sugar metabolism such as arabinose (MF0014), sucrose (MF0010), and melibiose (MF0009) degradation with higher BMI. There exists evidence from previous studies indicating that the gut microbiome may contribute to obesity development by increasing the energy extraction from food and how the body stores this energy^18^. In particular, several studies have pointed to groups of bacteria that may be more efficient at extracting energy from simple sugars and contribute to weight gain^17,18,32^. Our results also point to a pattern of higher BMI being associated with a higher abundance of genes related to energy extraction, supporting the idea that besides excessive calorie intake, obesity development is related to the ability of the gut microbiome to extract energy from food^18,32,33^.

Among other pathways associated with BMI (Table 3), we found propionate synthesis pathways (MF0095, MGB055, MF0094, and MGB054) negatively associated with BMI. Propionate is a type of short-chain fatty acid that is produced by certain types of bacteria in the gut. Increased gut propionate has been shown, including through targeted delivery studies, to reduce inflammation, improve insulin sensitivity and regulate appetite and body weight maintenance by promoting the secretion of Peptide YY (PYY) and Glucagon-like peptide-1 (GLP-1)^34,35^. Evidence of more than a decade of research has shown that gut microbiome alterations (both at taxa and functional levels) are associated with diet-induced obesity and are reversible, leading to the suggestion that weight loss programs targeting the gut microbiome can be used to treat obesity^3,8,36^. In this context, the evidence presented in this and our previous studies supports this premise. It highlights microbial taxa and their functions that may be targeted to improve the gut microbiome composition with concomitant positive effects in health outcomes, including obesity, cardiovascular health, mental health, and functional gastrointestinal disorders^23–26^.

Interestingly, 9 out of the 18 genera associated with BMI also changed their abundance in a pattern consistent with the beneficial effect of the weight loss program on the gut microbiome. In particular, their abundance is negatively associated with BMI, which increases at T2, or their abundance is positively associated with BMI and decreases at T2, highlighting these genera as attractive targets to understand how the microbiome may mediate the health benefits of weight loss. On the other hand, the nine genera we deemed not improved by the weight loss program warrant further research to identify food components, pre- and probiotics that could shift their abundance in a health-promoting direction to enable the lifestyle change to elicit additional health benefits. This reasoning stems from an expected linear relationship between the abundance and health benefits, which we understand is a valuable working model but limited due to the complex ecology of the gut microbiome where non-linear relationships are also expected.

Data analysis strategies considering each taxon need to account for or consider the community assembly in the gut microbiome. Our analyses of the predicted abundance of functional pathways at the community level help tackle this challenge but avoid exploiting the community assembly itself. To address this challenge, we carried out networks analyses to identify groups of taxa with tightly correlated abundance patterns that may reflect the underlying microbial community assembly (i.e., where nodes represent microbial taxa and edges represent their patterns of co-abundance) and are associated with potential consequences for human health^37–39^.

The weighted network analyses identified five network modules as the overarching gut microbiome structure among these individuals (Figure 4). Module 2 was significantly associated with BMI with a negative correlation (Table S5), and several of the genera on this module have been previously associated with BMI with the same directions of effect, including *Akkermansia*^40^, *Roseburia*^41,42^, *Butyricicoccus*^43,44^, *Lachnospira*^45^ and *Eubacterium ventriosum* group^46^. *Roseburia* and *Akkermansia* were significantly associated with BMI and changed their abundance between T1 and T2 (Table S3). Module 3 was significantly associated with BMI with a positive correlation and showed a significant decrease in abundance at time point T2. In line with our findings for module 2, several of the genera included in module 3 had been previously associated with BMI and individually were associated with BMI and changed their abundance between T1 and T2 (Table S3), including Oscillospiraceae^31^, Christensenellaceae^31^, *Alistipes*^4,44,47–49^ and *Sutterella*^50^. These findings support the notion that gut microbiome changes associated with obesity (or BMI), and dietary changes do not occur on individual taxa in isolation but on microbial communities whose members are dependent and complementary to each other regarding nutrient utilization, metabolite production, and niche occupation. These results warrant further research since these modules may provide an initial proposal to understand the microbiome community composition and dynamics regarding BMI and obesity across different individuals, genders, diets, and lifestyles.

Digbi’s dietary and lifestyle based weight loss program has been previously shown to be effective for weight loss. During this study’s microbiome sampling period (from T1 to T2), 17.5%, 31.1%, and 14.6% of individuals lost more than 3%, 5-10%, and more than 10% of body weight, respectively. A previous study showed that weight loss of 3%, 5%, and 10% has health economics implications with significant savings in total healthcare costs^51^. This suggests that the biomarkers identified are relevant not only to the understanding of obesity, and monitoring its prevention and reversal but also to monitoring the economic outcomes of weight loss programs.

This study has some limitations that are important to note. Firstly, the findings presented here are derived from a weight loss cohort and thus may not reflect the relationship between BMI and gut microbiome diversity and health in the broader population. Secondly, this observational study did not collect information regarding longitudinal changes in factors that influence the microbiome composition, such as disease diagnoses or measures of environmental, social, and work determinants of health. Thirdly, our associations with bacterial functional pathways relied on predicting the abundance of the relevant genes and did not directly relate to the activity at the enzymatic or molecular level. Fourthly, there were substantial variations in the time elapsed between T0 and T2 and T1 and T2. Despite not finding a relationship between these periods and the amount of weight loss, we cannot rule out a systematic effect on the changes in the microbiome observed. Fifth, Digbi Health weight loss program is an actively commercialized weight loss program. As a real-world data source, it cannot measure some variables that would generally be captured in research, like changes in medication intake, and significant changes in determinants of health, among others, that could have informed and explained some of the associations we have identified. Sixth, the cohort studied stems from a larger cohort and represents a subset that actively engaged in returning the two fecal samples for gut microbiome analyses and provided survey responses at T0, T1, and T2, including self-reported weight or engaged with Bluetooth weight scales. These previous limitations imply that our results may be affected by ascertainment or collider biases that we cannot effectively control due to this study’s observational and retrospective nature. Nonetheless, our findings provide new insights into the effects of dietary and lifestyle changes on the longitudinal evolution of the gut microbiome over time and its potential involvement in weight loss and warrant consideration by the broader scientific community.

## Methods

### Cohort enrollment and inclusion criteria

The study subjects enrolled in the Digbi Health personalized care program for weight loss, which is a commercially available product known as Digbi Control. Research associated with this weight loss program has been reviewed and approved by the Institutional Review Board of E&I Review Services (protocol code #18053 on 05/22/2018). All subjects considered for the present manuscript provided their research informed consent electronically as part of their written informed consent form.

For this retrospective study, the inclusion criteria were: 1) subjects with gut microbiome data at two-time points: T1: Early phase and T2: Follow-up phase; and 2) subjects did not self-report consuming antibiotics at the enrollment date.

### Digital therapeutics weight loss program

Digbi Health’s weight loss program has been described elsewhere^26^. In a nutshell, personalization of dietary plans is achieved by analyzing participants’ genetics, gut microbiome, lifestyle, and demographics. Based on these data, the program encourages participants to make incremental lifestyle changes focused on reducing sugar consumption and timing meals to optimize insulin sensitivity, reduce systemic inflammation by identifying possibly inflammatory and anti-inflammatory nutrients, and increase fiber diversity to improve gut health. Behavioral changes are implemented with the help of virtual health coaching and the app, ensuring they are habit-forming.

### Sample collection and processing

Subjects self-collected fecal samples using fecal swabs (Mawi Technologies iSWAB Microbiome collection kit, Model no. ISWAB-MBF-1200). Sample collection was completed by following standardized directions provided to all subjects in an instruction manual. DNA extraction was performed using Qiagen MagAttract Power Microbiome DNA Kit on an automated liquid handling DNA extraction instrument, followed by bacterial 16S rRNA gene V3-V4 region amplification and sequencing on the Illumina MiSeq platform using 2 × 300 bp paired-end sequencing performed at Akesogen Laboratories in Atlanta, GA. Sequence reads were demultiplexed, denoised and ASVs generated using DADA2 in QIIME2 version 2021.4^52^.

### Microbiome data analysis

We collected 206 stool swab samples (103 individuals at two-time points, T1: Early phase, and T2: Follow-up phase). Initial quality control steps included the removal of primers and low-quality bases, removing ASVs classified as non-bacterial sequences (such as Euryarchaeota, Chloroplast, Mitochondria), or unassigned taxa at the phylum level. Taxa were agglomerated at genus levels, and those with low abundance (taxa with <10 reads in at least 10% of samples) were excluded, resulting in a reduction of the sparsity of the abundance matrix from 99.75% to 37.6% (with an average of 98.3% of read retention) and removal of singletons. The abundance matrix was rarefied at even depth (n=61,000 reads per sample (minimum reads across the samples) with 500 iterations) using QIIME2^53^, resulting in 155 taxa across 206 samples. The abundance of functional microbial pathways related to gut and neuroactive metabolites^54^ was calculated with the q2-picrust2 plugin (v2.4.2) in QIIME2^55^ and the Omixer-RPM package (version 0.3.2)^56^. All raw abundances were centered-log ratio (CLR) transformed unless otherwise specified^57^.

### Network module analysis

We performed network analyses of the gut microbiome profiles to identify 1) network modules associated with BMI and BMI changes between the time points studied and 2) the key driving taxa of these network modules. We define network modules as a set of taxa with higher levels of correlation on their abundance among them compared to other taxa. We utilized the WGCNA software package for R^58^. We preprocessed the abundance profiles by calculating the CLR transformation and setting taxa with 0 counts as NA. We used the “pickSoftThreshold ‘‘ function to explore the relationship between the power parameter and the connectivity and selected power = 1 for the analyses. Higher values led to a drastic reduction of average connectivity, which strongly influenced the detection of modules which translated into detecting none or one with values of the power parameter >3. For module detection we used the “blockwiseModules’’ function with the following non-default parameters: power=1, corType=‘bicor’, corOptions=list(use=‘pairwise.complete.obs’, checkMissingData=TRUE, TOMType = “signed”, minModuleSize = 10, reassignThreshold = 0, mergeCutHeight = 0.25, numericLabels=TRUE, pamRespectsDendro=FALSE, and replaceMissingAdjacencies=TRUE). The per taxa missingness values filtered out 52 taxa with an excess of missing values and for which pairwise correlations with other taxa could not be calculated. Likewise, 1 sample was removed from the analysis due to 50% missing values. We used the “bicor” correlation due to the non-normal distribution of microbiome abundance values. We compared the results obtained using the “bicor” correlation with a signed or unsigned TOMType and found that the resulting modules were identical. The output of the “blockwiseModules’’ function provides an assignment of each taxon to a network module, a summary of the abundance patterns of the modules, and calculates the first eigenvector, which summarizes the main trend of the abundance matrix of the taxa associated with each module. We calculated the correlation between each taxa CLR value and the module eigenvector to identify the taxa that are key drivers of the network module as described by the software authors^59^.

### Statistical analysis

We used PERMANOVA, which performs community-level multivariate association with variables, based on the abundance matrix using the Bray-Curtis dissimilarity, using the vegan package (adonis2 function, strata= user.id) in R. Further to test for homogeneity of multivariate dispersions (comparing inter-individual variations) between groups, we used betadisper test. Comparisons of the diversity indices between time points (T1 vs. T2) were tested statistically using the Wilcoxon-signed rank test with FDR corrections.

To identify taxa and functional pathways associated with BMI and/or changes between T1 and T2, we used linear mixed models implemented on the GAMLSS software package for R^60,61^. In particular, we used the following regression formula “abundance ∼ 1 + Gender + Age + BMI + Time.point +re(random=∼1|User.id, method = ‘REML’)” with the following options ‘gamlss(…., control=gamlss.control(c.crit = 0.001, n.cyc = 200), family = BB())’, where “abundance” corresponds to the raw counts of the taxa or functions divided by the total counts of the sample and “BEZI()” is the zero-inflated beta, distribution family. We performed multiple corrections using the local FDR methodology implemented on the “ashr” software package for R^62^.

We tested the association between the network module’s eigenvector and BMI and time point using linear mixed models implemented on the lmerTest software package for R^63^. The BMI model used the following options “lmerTest::lmer(Module_EigenVector ∼ 1 + BMI + Gender + Age + (1|User.id), data=pheno_me))” for the time point model we replaced BMI for time point in the formula. We identified modules that were associated with either BMI or time point and those that are associated with both. Results were corrected for multi-testing using Sidak’s methodology, which calculates the family-wise corrected p-values *p*^*Sidaik*^ = 1 − (1 − *p*)^*k*^ where *p* is the univariate p-value and *k* = 5 is the number of hypotheses tested.

## Data Availability

The microbiome sequence data used in this study were submitted to NCBI SRA under Bioproject accession number PRJNA907500.

https://www.ncbi.nlm.nih.gov/bioproject/PRJNA907500

## Acknowledgments

We are grateful to the subjects participating in Digbi Health research studies without whom it would not have been possible to perform this research.

## Author contributions

SK and IP: formal analysis, methodology, software, visualization, writing – original draft, and writing – review and editing. BJ, KM: formal analysis software. SS, CI, formal analysis, methodology. PD and GK: conceptualization, and writing – review, and editing. DA: conceptualization, writing – original draft, and writing – review and editing. RS: conceptualization, funding acquisition, and writing – review and editing. All authors contributed to the article and approved the submitted version.

## Ethics declarations

Informed consent was obtained electronically from study participants. The study was conducted in accordance with the Declaration of Helsinki, and approved by the Institutional Review Board of E&I Review Services (protocol code #18053 on 05/22/2018).

## Supplementary information

### Supplementary Figures

**Figure S1.**
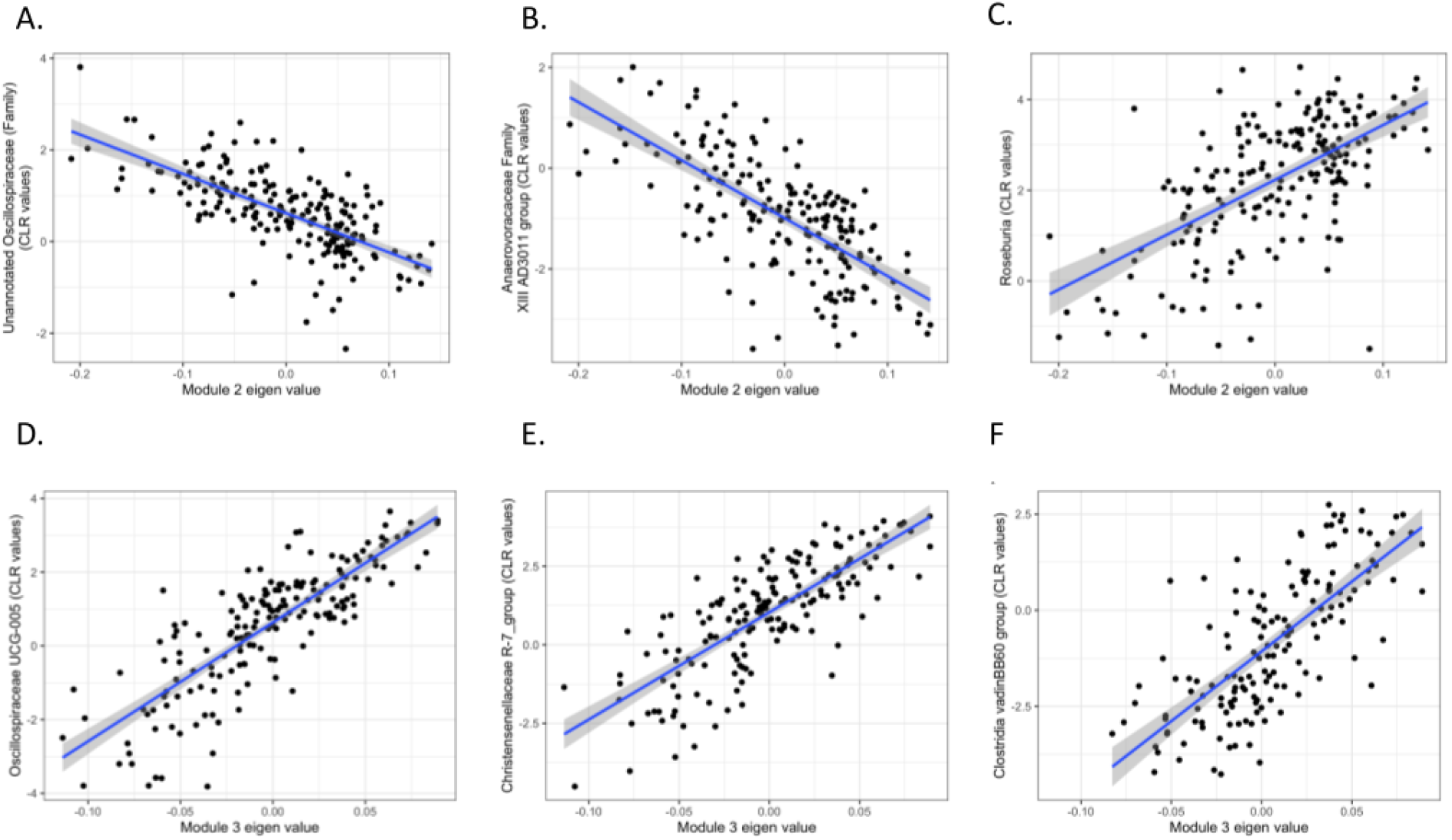
Association of key gut microbial genera and network modules’ eigenvectors. Correlation between module 2 (panels A, B, and C) and module 3 (panels D, E, and F) with the three genera with the most significant correlation with the corresponding eigenvector. (A) Anaerovoracaceae Family XIII AD3011 group, (B) Unannotated Oscillospiraceae (Family), (C) *Roseburia*, (D) Oscillospiraceae UCG-005, (E) Christensenellaceae R-7 group, and (F) Clostridia vadinBB60 group.

### Supplementary Table Legends

**Table S1. Effect of demographic and lifestyle factors on weight loss during the program**.

**Table S2. Effect of demographic, lifestyle and BMI factors on gut microbiome alpha diversity measures**.

**Table S3. Results from the Linear Mixed Models (GAMLSS BEZI) based regression analysis between taxa and variables that were seen associated with overall gut microbial diversity**.

Taxa highlighted in green are the ones with significant association with the respective variable (multiple corrections using the local FDR methodology).

**Table S4. Results from the Linear Mixed Models (GAMLSS BEZI) based regression analysis between microbial pathways and variables that were seen associated with overall gut microbial diversity**.

Microbial pathways highlighted in green are the ones with significant association with the respective variable (multiple corrections using the local FDR methodology).

**Table S5. Network modules 2 and 3 and the correlation statistics of each genera with the module eigenvector**.

**Table S6. Demographic and clinical characteristics of participants enrolled in the study**.

